# How rare and common risk variation jointly affect liability for autism spectrum disorder

**DOI:** 10.1101/2020.10.27.20220095

**Authors:** Lambertus Klei, Lora Lee McClain, Behrang Mahjani, Klea Panayidou, Silvia De Rubeis, Anna-Carin Säll Grahnat, Gun Karlsson, Yangyi Lu, Nadine Melhem, Xinyi Xu, Abraham Reichenberg, Sven Sandin, Christina M. Hultman, Joseph D. Buxbaum, Kathryn Roeder, Bernie Devlin

## Abstract

**Background:** Genetic studies have implicated rare and common variation in liability for autism spectrum disorder (ASD). Of the discovered risk variants, those rare in the population invariably have large impact on liability, while common variants have small effects. Yet, collectively, common risk variants account for the majority of population-level variability. How these rare and common risk variants jointly affect liability for individuals requires further study.

**Methods:** To explore how common and rare variants jointly affect liability, we assessed two cohorts of ASD families characterized for rare and common genetic variation (Simons Simplex Collection and Population-Based Autism Genetics & Environment Study). We analyzed data from 3,011 affected subjects, as well as two cohorts of unaffected individuals characterized for common genetic variation: 3,011 subjects matched for ancestry to ASD subjects; and 11,950 subjects for estimating allele frequencies. We used genetic scores, which assessed the relative burden of common genetic variation affecting risk for ASD (henceforth burden), and determined how this burden was distributed among three subpopulations: ASD subjects who carry a rare damaging variant implicated in risk for ASD (mutation carriers); ASD subjects who do not (non-carriers); and unaffected subjects, who are assumed to be non-carriers.

**Results:** Burden harbored by ASD subjects is stochastically greater than that harbored by control subjects. For mutation carriers, their average burden is intermediate between non-carrier ASD and control subjects. Both carrier and non-carrier ASD subjects have greater burden, on average, than control subjects. The effects of common and rare variants likely combine additively to determine individual-level liability.

**Limitations:** Only 258 ASD subjects were known mutation carriers. This relatively small subpopulation limits this study to characterizing general patterns of burden, as opposed to effects of specific mutations or genes. Also, a small fraction of subjects that are categorized as non-carriers could be mutation carriers.

**Conclusions:** Liability arising from common and rare risk variation likely combine additively to determine risk for any individual diagnosed with ASD. On average, ASD subjects carry a substantial burden of common risk variation, even if they also carry a rare mutation affecting risk.

## Background

The genetic architecture of autism spectrum disorder (ASD) remains uncertain, although there are putative models(1). One model posits that ASD is heterogeneous, that severe mutations in any of a large set of genes are sufficient to cause the disorder. Another emphasizes the major role played by common variation – shared by all of us to a greater or lesser extent – in the documented high heritability of ASD(2-6). A hybrid model asserts that common and rare variation combine in some way, perhaps additively(7), to confer liability(1, 7, 8). At the level of a population, common variation probably plays the dominant role in liability, whereas a rare mutation can make the largest contribution to liability for an individual who carries it(9). Still, our current understanding of the genetic architecture of ASD is unsatisfactory, especially regarding how common and rare variation jointly confer risk. This architecture is important because it has clinical consequences; for example, it could require more nuanced evaluation of recurrence risk. Establishing the exact nature of the interplay between common and rare risk variation will be challenging, however, because of the multiplicity of plausible models that could fit the current data.

To address this problem empirically, a sample of ASD subjects who have been characterized for rare and common variation is essential. Because rare, *de novo* damaging mutations carry the most readily detectable signal for ASD association(10-13), the ideal sample would be characterized for such mutations. Following the tradition in human genetics, we call ASD subjects carrying such damaging mutations as “mutation carriers” and all other ASD subjects as “non-carriers”. Such well characterized samples of the population are not common and none as yet are especially large, a limiting factor for any study. For the sample to be analyzed here, we combine data from three sources: subjects diagnosed with ASD from the Simons Simplex Collection or SSC(14); ASD and unaffected subjects from the Population-Based Autism Genetics & Environment Study or PAGES(9); and subjects from the Electronic MEdical Records and Genomics Network or eMERGE(15), whom we assume have not been diagnosed with ASD and are non-carriers.

These data could be analyzed in at least two ways. One approach would be to use polygenic risk scores (PRS), which are based on common variants putatively affecting liability(16). Typically, these variants are identified from genomewide association studies (GWAS) and the PRS for each subject is computed as a weighted sum of the count of risk alleles they carry. Then, values of the PRS in ASD mutation carriers and ASD non-carriers, as well as unaffected subjects, can be contrasted to assess how common and rare variation jointly confer risk. An elegant version of this approach is the pTDT or polygenic Transmission Distortion Test(7), which requires parental genotypes. Because only a portion of our data have parental genotypes, here we concentrate on the PRS.

The PRS is only as effective as the information arising from GWAS, which for ASD is still relatively limited compared to other phenotypes (see Grove and colleagues(6) versus the Psychiatric Genomics Consortium for schizophrenia and bipolar disorder (17, 18)). For this reason, we emphasize here another approach to developing a score, which will be based on the theory of Genomic-Best Linear Unbiased Prediction (G-BLUP)(19-21). The ideas behind G-BLUP are similar to the PRS. Rather than using GWAS results, G-BLUP develops a predictive model to distinguish case versus control subjects, genetically, using genetic variation across the genome. Here we call this genomic prediction “GP” and show how to produce effective and unbiased estimates in the context of ASD, although the approach we lay out applies to any phenotype.

While the PRS and GP are not strictly independent, they can be combined to produce an even more effective predictor. We use these approaches to (1) document that the burden of risk variants carried by ASD subjects is stochastically greater than that carried by control subjects; (2) that carriers of rare, damaging mutations bear a burden intermediate between non-carrier and control subjects; (3) that both mutation carriers and non-carriers have a stochastically greater burden of common risk variation than control subjects; and (4) that the effects of common and rare variants on liability for ASD likely combine additively. Regarding (3), it appears that ASD subjects carry a substantial burden of common risk variation, even if they also carry a rare mutation affecting risk. For (4), although common and rare risk variation likely act additively, the resolution imposed by current data is coarse.

## Methods and Results

### Data

Here we present analyses of 17,972 samples of European descent (Additional file 1: Fig. 1): 3,011 were diagnosed with ASD; while the remaining 14,961 were assumed to be unaffected. Among the ASD subjects, 1,996 and 1,015 came from SSC and PAGES, respectively. Control subjects came from the PAGES (1,524) and eMERGE (13,437) cohorts. All subjects from the SSC and eMERGE were collected in the USA; subjects for PAGES come from Sweden. DNA from all subjects was genotyped on an Illumina genotyping platform: Human1M_v1, Human1M_Duov3, and HumanOmni-2.5 for SSC; Infinium OmniExpressExome-8 V1, Infinium OmniExpressExome-8 V1.1-V1.4 for PAGES; and Illumina Human660W_Quad_v1_A for eMERGE. Genotypes for all samples were imputed at the Michigan Imputation Server(22) using the HRC reference panel(23). Post-imputation quality control reduced the number of SNPs to 5,145,175. SNPs in high linkage disequilibrium (LD) were thinned using PLINK 2.0(24) (--clump-r2 0.81; --clump-kb 50), producing 910,356 SNPs; SNPs that were physically genotyped in more subsets were given preference in the thinning process. A final pruning step with parameters r^2^ < 0.64 for moving blocks of 50 SNPs and a window of 5 SNPs at a time reduced the genotyped dataset to 553,406 SNPs to be used for all GP calculations.

**Figure 1.**
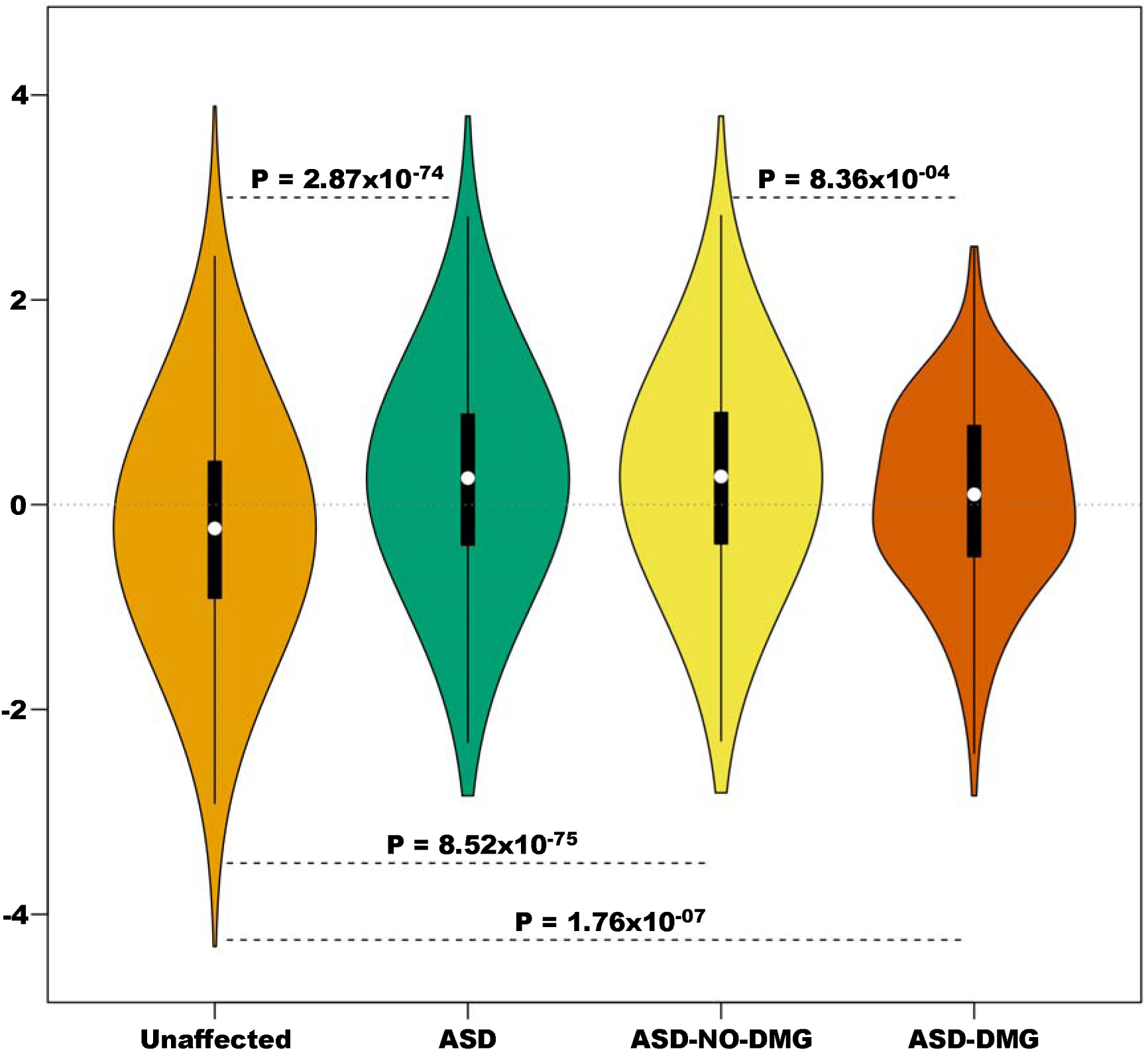
The burden of risk variation by ASD status and by carrier status of ASD subjects. Burden is estimated using GP, standardized to have mean=0 and standard deviation=1. ASD-NO-DMG, ASD subjects with no known damaging variants; ASD-DMG, ASD subjects with known damaging variants.

### Effective GP estimates

#### Overview

Our goal in this section is to develop a G-BLUP estimation procedure for GP that distinguishes ASD from unaffected subjects and is free of the influence of ancestry. Note that G-BLUP relies on genetically estimated relationships among subjects in a sample. To calculate this genomic relationship matrix (GRM), genotypes are standardized according to allele frequencies of the SNPs(25, 26). Data from multiple subpopulations can be challenging if the number of subjects per subpopulation is unbalanced because allele frequency estimates are dominated by the largest subpopulation(s). As a result, the GRM is well estimated within the main subpopulation, but relationship estimates for subjects in other subpopulations will tend to be biased. To overcome this concern, we divide our data into ancestry clusters; determine allele frequencies within clusters; standardize the genotypes within clusters; and then calculate the GRM based on these cluster-specific standardizations. This is not the sole concern regarding experimental design. If the ASD and unaffected subjects are not balanced over the ancestry space, bias can be induced in estimation of GP. Specifically, this imbalance can produce misleading differentiation between ASD and unaffected subjects. For this reason, we estimate GP based on a set of ancestry-matched case-control samples(27), thereby avoiding the problem of unbalanced sampling.

#### Ancestry

To assess ancestry, 99,509 SNPs were selected because they were genotyped for all samples and were relatively independent (the larger set of SNPs was pruned to reduce linkage disequilibrium, as measured by pairwise r^2^: 50 SNP moving blocks, 5 SNP at a time, pairwise r^2^ <0.64). Using genotypes from these SNPs, genetic ancestry was determined using function ‘clusterGem’ in the package GemTools(28). All 17,972 samples were clustered simultaneously using three ancestry eigenvectors, which divided the sample into four clusters (Table 1, Additional file 1: Fig. 1). Within each cluster, genetically matched pairs were chosen using the function ‘pairmatch’ in library *optmatch* in R (1-to-1 fullmatch), which assessed pairwise distances among subjects based on the space defined by three ancestry eigenvectors.

**Table 1.** Distribution of the subjects over the four ancestry clusters by ASD status and cohort.

| Cluster | ASD | Unaffected | Total | ASD |  | Unaffected |  |
| --- | --- | --- | --- | --- | --- | --- | --- |
|  |  |  |  | SSC | PAGES | PAGES | eMERGE |
| CL1 | 626 | 4034 | 4660 | 625 | 1 | 10 | 4024 |
| CL2 | 873 | 5407 | 6380 | 813 | 60 | 74 | 5433 |
| CL3 | 458 | 976 | 1434 | 372 | 86 | 165 | 811 |
| CL4 | 1,054 | 4444 | 5498 | 186 | 868 | 1275 | 3169 |
| Total | 3011 | 14961 | 17972 | 1996 | 1015 | 1524 | 13437 |

#### Identifying carriers of damaging mutations

ASD subjects were classified as “mutation carriers” of a damaging mutation if their DNA contained a pathogenic CNV or had a *de novo* protein truncating variant (PTV) or deleterious missense variant (Missense badness, PolyPhen-2, and Constraint score MPC>2)(29) in one of 102 genes identified in Satterstrom et al.(10) as affecting risk. For the PAGES sample, we used the set of pathogenic CNVs described in Mahjani et al. (in review), who identified CNVs from 956 out of the 1015 AD subjects we analyzed. For the SSC sample, CNVs were identified by Sanders et al(11) and pathogenicity defined following Mahjani et al. (in review). For each dataset, subjects who carried a trisomy or had large or multiple CNVs were set to “undetermined” for carrier status, thus were not in the mutation carrier versus non-carrier analyses, although they were retained as ASD subjects.

DNA from ASD subjects from the SSC sample was characterized for PTV and missense (MIS) variants using whole-genome sequence, as reported by An et. al(30); DNA from PAGES ASD subjects was characterized for PTV and MIS variants from whole-exome sequence, as reported in Satterstrom et al.(10) (778 out of 1,015 AD subjects we analyzed). For the PAGES subjects whose DNA was not characterized, we assumed they were non-carriers.

DNA from 305 ASD subjects carried one or more damaging mutations (Table 2, Additional file 1: Table 1). If a subject carried multiple damaging mutations, the most severe mutation for each subject was counted, under the assumption that the ranking of severity was CNV > PTV > MIS.

**Table 2.** Counts of most severe damaging mutation (CNV > PTV > MIS).

|  | **Total** | **SSC** | **PAGES** |
| --- | --- | --- | --- |
| CNV | 166 | 78 | 88 |
| PTV | 78 | 58 | 20 |
| MIS | 61 | 38 | 23 |
| Non-carrier | 2682 | 1,814 | 868 |

#### Genomic Relationship Matrix

Using unaffected subjects *not matched* to ASD subjects, we obtained within-cluster and overall allele frequency estimates. Then, from these two estimates, we used empirical Bayes methods, as described in Bodea et al.(31), to determine final cluster-specific allele-frequency estimates. These frequencies were then used to standardize cluster-specific genotypes and compute a cluster-specific GRM. We call this the cluster-specific GRM, or CLS-GRM, to differentiate it from a GRM computed from genotypes standardized by the mean allele frequency for each SNP using all 11,950 unmatched controls, which we call POP-GRM. Finally, we computed a GRM using the default approach implemented in GCTA software(25, 26), which uses all 17,972 samples (GCTA-GRM). For the GCTA-GRM, rather than matching, we follow tradition by computing the first 10 principal components of ancestry, using GCTA software, and using them to account for ancestry in GP calculations.

#### Approach to G-BLUP estimation of GP

For GP to discriminate ASD from unaffected subjects, the expected number of risk alleles in ASD subjects should be stochastically greater than that in unaffected subjects. As the difference in the relative number of risk alleles, which we call the “burden”, becomes greater between the two subpopulations, the GP becomes more accurate. A common way to build a GP is to split the data into a training and a test set. For the training set, diagnosis is known and is used to develop the model for GP, which is then evaluated in the test set. A common breakdown is to use 90% of the sample for training, which can be done repeatedly using different portions of the sample. An expectation of statistical theory is that a larger training set yields a more accurate GP. For this reason, we chose a training and testing plan with N-2 observations for training and a matched pair for testing. This is iterated over all matched possible pairs, making it also a computationally intensive plan. To make the plan feasible, we implemented computing techniques to expedite calculations, as described in Additional File 1.

We analyzed N=3011 matched pairs. CLS-GRM was used for the genetic relationship among the matched samples, without additional covariates. G-BLUP calculations require an estimate of heritability for ASD, which we set at 0.70(32). GP estimates were standardized to have mean=0 and standard deviation=1.

#### Results for GP

To ensure matching was adequate, we first tested whether GP differed between clusters, after controlling for diagnosis. In this analysis-of-variance model, GP did not differ significantly by cluster (F=0.795, df = 3, 6017; p = 0.497). However, GP was significantly different (p = 7.61⨯10^−81^) between ASD and unaffected subjects (Fig. 1), showing that the average burden of risk variants was greater in ASD subjects. The relative risk of being an ASD subject, as opposed to unaffected, also increased with GP (logistic regression OR = 1.67; 95%CI = 1.58-1.77; P = 2.87⨯10^−74^; pseudo-R^2^ = 7.80%). Because GP is continuous and standardized, the increased risk should be interpreted in units of standard deviations of GP. Within ASD subjects, mutation carriers had a significantly smaller burden of common risk variants, on average, than do non-carriers (Fig. 1), which is also reflected in the relative risk as a function of GP (OR = 0.81; 95%CI = 0.71-0.92; P = 8.36⨯10^−4^). Both mutation carriers and non-carrier ASD subjects had a greater average GP than unaffected subjects (Fig. 1). Including 3 or 10 eigenvectors of “ancestry” as covariates in the model did not alter these conclusions (Additional file 1: Table 2), although adding 10 eigenvectors as covariates diminished somewhat the difference in GP between ASD and unaffected subjects, showing that eigenvectors can be a function of both the burden of risk variation and ancestry.

Conducting other exploratory analyses, we showed the following: CLS allele frequencies were better for standardizing genotypes than population-level allele frequencies (Additional file 1: Table 3); in some settings, adjusting by eigenvectors of ancestry can overcome the inaccuracy induced by population-level allele frequencies (Additional file 1: Table 4); our training/testing plan of N-2/2, where the 2 individuals comprise a matched pair, was optimal relative to other N-X/X for X > 2 (Additional file 1: Table 5); and, for data sets imbalanced in affected and unaffected subjects within the genetic ancestry space, such as Table 1, this imbalance biased GP (Additional file 1: Fig. 2).

**Table 3.** Logistic regression of ASD status on G-BLUP and DMG status in cases on G-BLUP, ASD-PRS, SCZ-PRS, and the weighted genomic risk score (WGRS).

| Score | ASD status |  |  |  | DMG stats (cases only) |  |  |
| --- | --- | --- | --- | --- | --- | --- | --- |
|  | OR | 95%CI | P | Pseudo-R<sup>2</sup> (%) | OR | 95%CI | P |
| G-BLUP | 1.67 | 1.58-1.77 | $2.87 \times 10^{-74}$ | 7.80 | 0.81 | 0.71-0.92 | $8.36 \times 10^{-4}$ |
| ASD-PRS | 1.21 | 1.15-1.28 | $1.28 \times 10^{-13}$ | 1.23 | 0.79 | 0.70-0.89 | $9.29 \times 10^{-5}$ |
| SCZ-PRS | 1.19 | 1.13-1.25 | $6.77 \times 10^{-11}$ | 0.95 | 0.89 | 0.79-1.00 | 0.0445 |
| WGRS | 1.73 | 1.64-1.83 | $1.56 \times 10^{-82}$ | 8.59 | 0.77 | 0.68-0.87 | $4.38 \times 10^{-5}$ |

**Figure 2.**
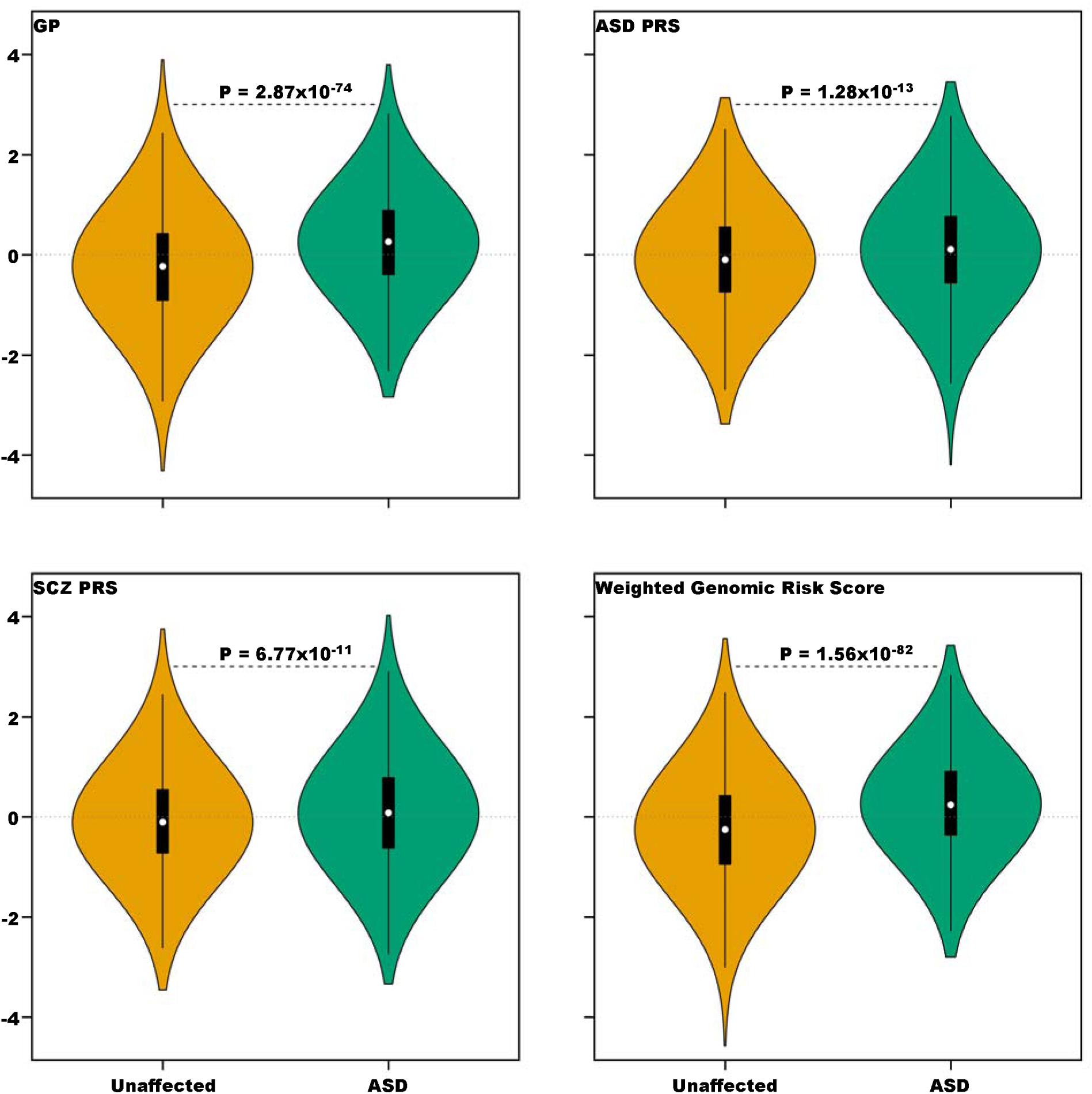
Distribution of risk scores divided by ASD and unaffected subjects. Burden is estimated using GP, an ASD polygenic risk score (ASD PRS), a schizophrenia polygenic risk score (SCZ PRS), and a weighted genomic risk score that incorporates information from GP, ASD PRS, and SCZ PRS.

### Polygenic Risk Scores and a weighted score

#### Approach

We evaluated two PRS, specifically one derived from an ASD GWAS(6) and the other from a recent schizophrenia (SCZ) GWAS(17). For the ASD GWAS, we used the results based only on the Danish iPsych data because SSC and a portion of the PAGES data were included in the full GWAS analysis(6). After quality control (QC), described in the Additional file 1 and using a GWAS threshold of p < 0.01, 9,983 and 26,972 SNPs were included for ASD and SCZ PRS, respectively.

#### Results for PRS

For the 3011 matched pairs previously described, both the ASD-PRS and the SCZ-PRS distinguished ASD from unaffected subjects, on average (Table 3, Fig. 2). The ASD-PRS also was significantly different for carrier versus non-carrier ASD subjects, although the SCZ -PRS is not (Table 3, Fig. 3).

**Figure 3.**
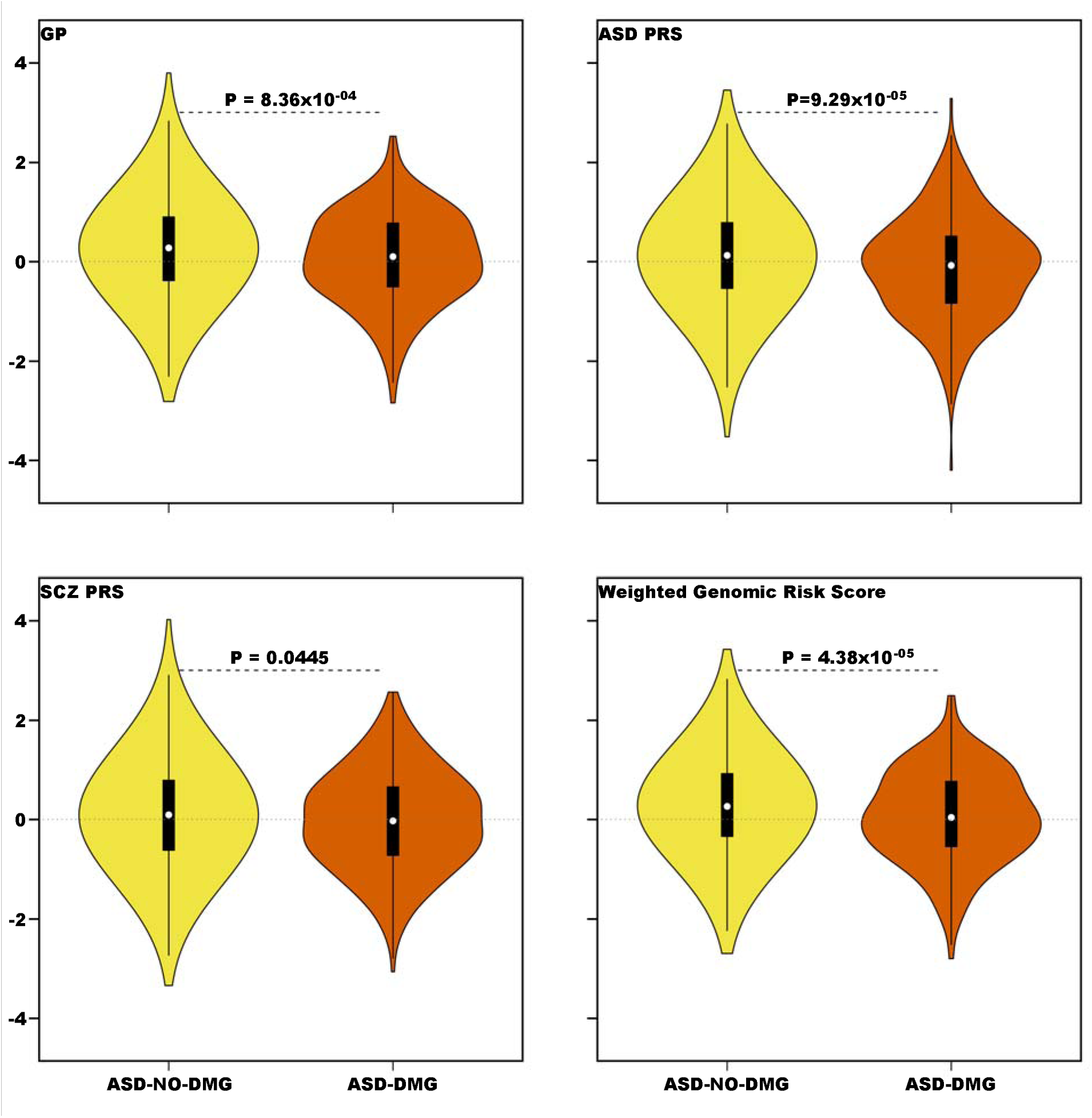
Distribution of risk scores divided by ASD mutation carriers and non-carriers. Burden is estimated as noted in Figure 2, here comparing ASD mutations carriers (ASD-DMG) to non-carriers (ASD-NO-DMG).

#### Combining scores

The correlation between GP and ASD-PRS was 0.079 (p = 9.80⨯10^−10^), while it was 0.099 for GP and SCZ-PRS (p = 1.16⨯10^−14^). ASD-PRS and SCZ-PRS were stochastically uncorrelated (r = 0.007, P = 0.581). Thus, given their modest correlations, these three scores had essentially independent information. To combine them into one weighted genomic risk score (WGRS), we based the weights on their Nagelkerke’s pseudo-R^2^, which roughly measures each score’s ability to separate ASD and unaffected subjects (Table 3). The WGRS was standardized to have mean 0 and standard deviation of 1. Compared to the other scores, WGRS better distinguished ASD from unaffected subjects and carrier/non-carrier status of ASD subjects (Table 3, Figure 2-3).

### Relationship between common and rare risk variants

We next ask about the interplay of common and rare risk variants. While diagnosis of ASD is binary, a person either does or does not meet diagnostic criteria, continuous variability of phenotypes related to ASD has long been recognized. A related mathematical observation is that a continuous liability model, in which a normally distributed liability is determined by genetic and environmental risk factors carried by each subject in a population, can be an excellent model for a binary trait like ASD (Fig. 4). In such a model, a liability threshold *t* determines whether an individual meets the diagnostic criteria and this threshold maps onto ASD prevalence (Fig. 4). If we take prevalence to be 1.5% for the population(10), it sets the threshold for diagnosis of ASD, in terms of a z-score, establishes the mean liability for ASD and control subjects, and thus defines the difference in average liability between ASD and unaffected subjects (Fig. 4).

**Figure 4.**
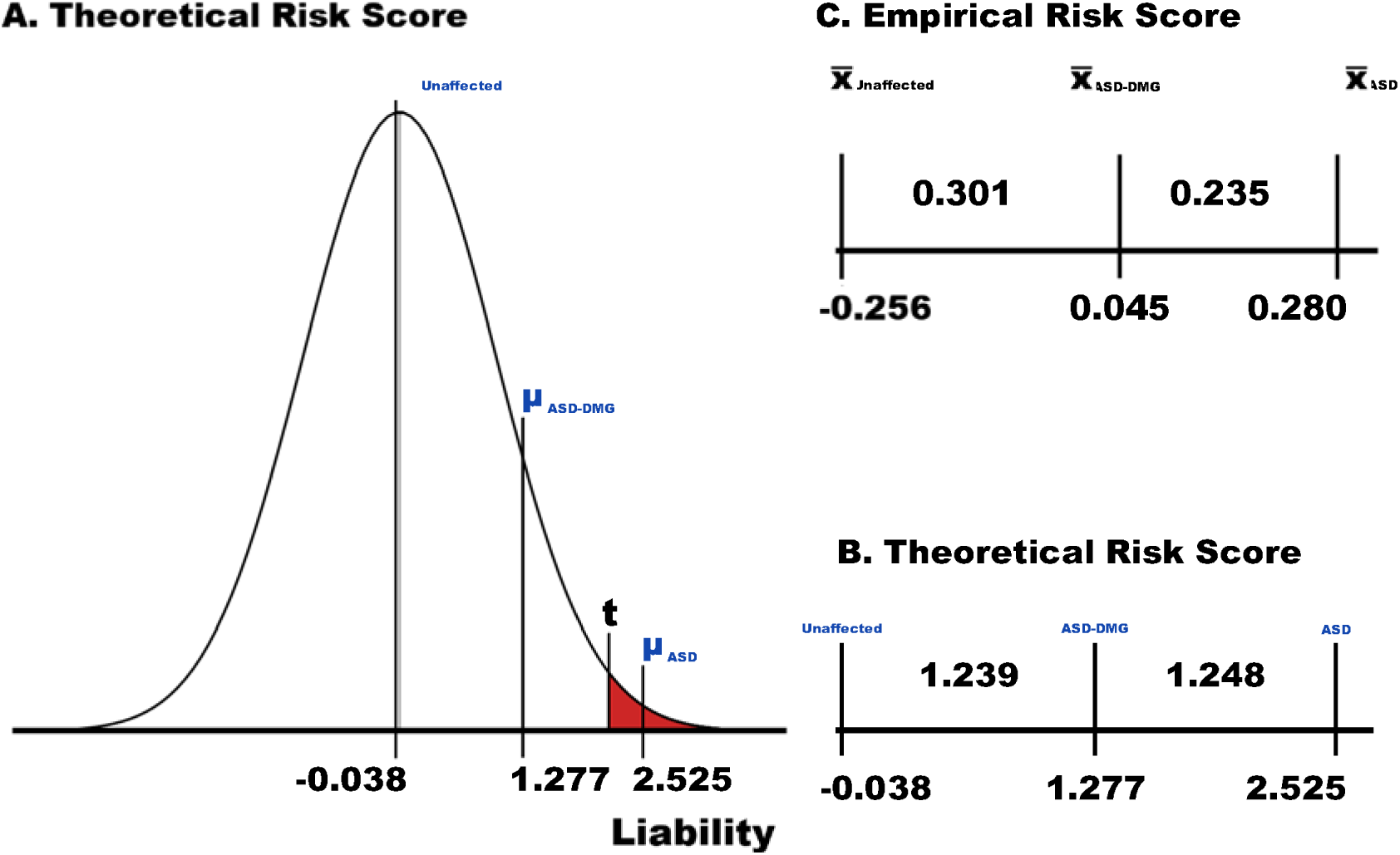
Continuous liability model for ASD compared to empirical realization. A. Liability is assumed to be normally distributed in the population, and there exists a threshold t of liability beyond which everyone is affected and below which no one is affected. ASD prevalence determines t and the average risk for ASD and unaffected subjects; given an estimate of the relative risk for ASD due to rare damaging mutations, the average risk for mutation carriers of these mutations is also specified. B. A portion of A, highlighting the relatively even spacing between its average risks. C. The realized average risk for these three groups, as measured by the weighted genomic risk score (Fig. 3).

For this prevalence, the average liability for ASD subjects is 2.525 and for unaffected subjects it is slightly below zero, -0.038 (Fig. 4A). To determine where the average carrier ASD subject would fall on this continuum of liability, we require an estimate of the relative risk due to such mutations. Using results from Satterstrom et al.(10) and Sanders et al.(11), a relative risk of 15 is a good approximation. Then, using standard theory described in Satterstrom et al.(10), liability for the average carrier would be 1.277 (Fig. 4A), which falls close to the mid-point between the average liability of ASD and unaffected subjects, 1.282 (Fig. 4A, B). How does this compare to results for WGRS (Fig. 4C)? For ASD mutation carriers, the average WGRS is 0.045, close to the midpoint (0.012) between the average WGRS for ASD non-carriers (0.280) and unaffected subjects (−0.256). Thus, these calculations show that rare damaging risk mutations and common risk variation act roughly additively to confer liability to ASD.

The number of ASD mutation carriers is too few to evaluate liability much more deeply, at least reliably. If the additive model were a close approximation to reality, we would anticipate that the burden observed in ASD subjects would vary inversely with the relative risk for ASD associated with the damaging rare mutations they carry. Estimated relative risks associated with pathogenic CNVs and LoF tend to be similar and large(11), whereas the relative risk associated with missense mutations tend to be far smaller. When we partition mutation carriers according to the type of mutation they harbor and compare their burden to that of non-carriers and unaffected subjects, results are consistent with an additive model (Fig. 5): the average burden for ASD subjects carrying CNVs is smallest, although only slightly smaller than the average burden of those carrying LoF mutations; as might be expected, the difference between these two groups is not meaningful; whereas the average burden of ASD subjects carrying missense variants is substantially larger. By contrast, one might expect that carriers with mutations in genes commonly disrupted in ASD subjects, such as *CHD8*, would bear a smaller burden, on average, than carriers of mutations in genes disrupted far less often. A confounder here is gene size, larger ASD risk genes will tend accrue more mutations than smaller genes. While imperfect, a rough way to account for this confounder is to use the TADA Bayes factor(33), which summarizes the evidence for association from the expected spectrum of mutations, based on gene size and nucleotide content, to that observed in the sample. Comparing this Bayes factor ranking for the 102 inferred ASD genes identified in Satterstrom et al.(10) to the burden harbored by ASD subjects carrying mutations in those genes, we do not find a significant relationship (Spearman rank correlation = -0.150, p=0.073), although the correlation is negative, as expected.

**Figure 5.**
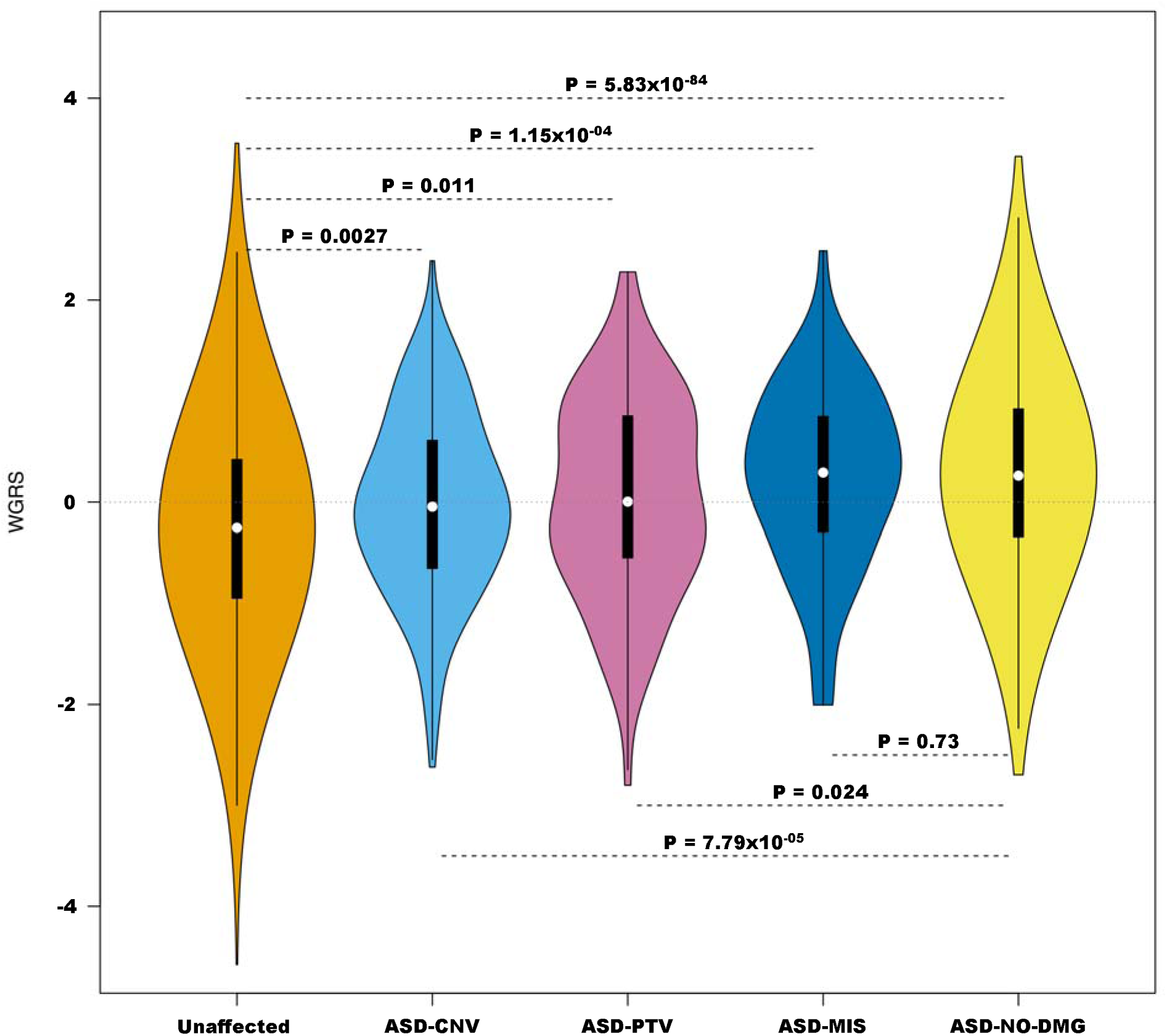
Distribution of risk scores for different subsets of subjects. Controls (unaffected), ASD mutation carriers with copy number variants (ASD-CNV), loss of function variants (ASD-PTV), and missense variants (ASD-MIS) and ASD non-carriers (ASD-NO-DMG).

## Discussion

Here we asked how rare and common risk variation jointly affect liability for ASD. We analyzed two samples characterized for both types of variation. Based on genotypes of common variation, we computed a small set of risk scores, each of which is likely to describe a portion of the genetic risk for ASD attributable to common variation. We also computed a weighted average of these scores, WGRS, which tended to perform better than any single score at differentiating ASD and unaffected subjects (Fig. 3) and at differentiating ASD subjects who carried rare mutations likely to affect risk – mutation carriers (Fig. 4) – from ASD subject who were not known to carry such variants (non-carriers). By contrasting patterns of the expected and observed burden of common risk variation in mutation carriers and non-carriers (Fig. 3-4), we conclude that the preponderance of evidence suggests that rare and common risk variation combine additively in their effects on ASD liability. This agrees with conclusions from other researchers(7, 34).

It is worthwhile emphasizing, however, that previous studies did not rigorously evaluate additivity and even the evidence presented here is far from conclusive. With larger samples, more compelling evidence could be drawn from an evaluation of carriers of mutations in genes with very different recurrence rates in ASD individuals. For example, certain genes, such as *CHD8*, are often found to carry mutations in ASD individuals. Other genes show significant association, yet far less recurrence. In future studies and with a much larger sample, we should be able to order ASD risk genes accurately in terms of the relative risk for ASD generated by mutations in these genes and evaluate how common variant risk changes along this ranking. If the two sources of risk work additively, they should show a strong negative relationship.

Why do we need to evaluate the nature of the relationship between common and rare variation so thoroughly? Suppose, for example, that some of the ASC’s 102 ASD genes do not truly affect risk and half of the *assumed* mutation carriers have mutations in these genes. Under this unlikely but not impossible scenario, these subjects would, in expectation, carry the mean WGRS observed in non-carriers (Fig. 4). To achieve the mean WGRS observed for the entire population of *assumed* mutation carriers, which consists of an equal mixture of true carriers of risk mutations and non-carriers, the mean for the subpopulation of true carriers would, in expectation, fall at the mean for unaffected individuals (Fig. 4). Under this scenario, joint effects of rare and common risk variants are irrelevant, a damaging rare variant would always be sufficient to cause ASD. Such scenarios can only be completely ruled out by using alternative ways of evaluating whether rare and common risk variation combine additively in their effects on ASD liability.

If common variant risk burden of mutation carriers is substantial, as the results here suggest, they have implications for genetic counseling regarding recurrence risk for ASD. Currently, genetic counseling for recurrence risk is binary, depending on whether or not a rare damaging mutation in an ASD gene is found in the proband’s genome. If such a mutation is found, then the mutation is typically assumed “causal’ for the proband’s ASD and recurrence probability for ASD is its prevalence. When this assumption is a good approximation, and it will be for many mutation carriers(9), counseling is also a good approximation. In some families, however, the mutation carrier has ASD in large part because of the polygenic burden carried by the parents and in this instance the current advice for recurrence risk is inaccurate. To give a concrete example, when we examined loss-of-function carriers in the SSC, we estimated that over 40% of these individuals would still have ASD even without the loss-of-function mutation(9), and this would be predicted to be even more of an issue with less penetrant variation (e.g., missense variation). Because the present state of knowledge does not allow us to know, a priori, which scenario is relevant, it is important for genetic counselors to consider this uncertainty and whether it should be built into their advice for parents regarding recurrence risk.

## Limitations

An ideal study would have even larger sample size than the one we present here. Given the incomplete knowledge of genes and mutations in ASD, a small fraction of subjects that are categorized as non-carriers could be mutation carriers. In addition, not all of DNA from PAGES ASD subjects was characterized for rare sequence variants through exome sequencing and they were assumed to be non-carriers because the vast majority would be.. These drawbacks limit our ability to go beyond coarse characterization of interplay of rare and common variation.

An ideal study would have a larger sample than the one in the study we present. Given the incomplete knowledge of genes and mutations in ASD, a small fraction of subjects that are categorized as non-carriers could be mutation carriers. In addition, not all of DNA from PAGES ASD subjects were characterized for rare sequence variants through exome sequencing and they were assumed to be non-carriers because the vast majority would be. These drawbacks limit our ability to go beyond coarse characterization of interplay of rare and common variation.

## Conclusion

While rare and common variation confer liability for ASD, how they jointly confer liability is an open question. By analyzing data from 3,011 affected subjects and 3,011 genetically-matched unaffected subjects, we conclude that the burden of common risk variants borne by ASD subjects is stochastically greater than that borne by control subjects; and that ASD subjects who carry rare damaging variation conferring risk for ASD have an average burden intermediate between non-carrier ASD and control subjects. The effects of common and rare variants likely combine additively to determine individual-level liability.

## Supporting information

AdditionalFile.1

## Data Availability

All rare variant data used in this manuscript are available in NIH- or SFARI-controlled data sets. Common genetic variation has been used by and is available via the Psychiatric Genomics Consortium (PAGES) or from SFARI.

## Abbreviations

AD: autistic disorder
ASD: Autism spectrum disorder
CLS: cluster-specific
CNV: copy number variant
eMERGE: Electronic MEdical Records and Genomics Network
G-BLUP: Genomic-Best Linear Unbiased Prediction
GCTA: Genome-wide Complex Trait Analysis
GP: genomic prediction
GRM: genomic relationship matrix
GWAS: genomewide association studies
HRC: Haplotype Reference Consortium
LD: linkage disequilibrium
MIS: *de novo* missense variant
PAGES: Population-Based Autism Genetics & Environment Study
POP: population
PRS: polygenic risk score
pTDT: polygenic Transmission
PTV: Protein truncating variant
QC: quality control
SSC: Simons Simplex Collection
SCZ: schizophrenia
WGRS: weighted genomic risk score

## Declarations

### Ethics approval and consent to participate

Not applicable.

### Consent for publication

Not applicable.

### Availability of data and material

All results from analyses are presented in the manuscript and Additional File 1.

### Authors’ contributions

L.K., K.P., S.DR., N.M., A.R., S.S., C.M.H., J.D.B., K.R., and B.D. conceived the study; A.C.S.G. and G.K. recruited subjects, with guidance from A.R., S.S., C.M.H., and J.D.B.; L.K., L.M., B.M., K.P., Y.L., and X.X. performed computational analyses with input from J.D.B., K.R., and B.D; L.K., L.M., B.M., S.DR., S.C., A.R., S.S., C.M.H., J.D.B., K.R., and B.D. wrote the manuscript. All authors approved of the final manuscript.

## Acknowledgements

We thank everyone who contributed to this study, both the research subjects and the investigators of the following studies:

## Simons Simplex Collection

We would like to thank the SSC principal investigators (A. L. Beaudet, R. Bernier, J. Constantino, E. H. Cook, Jr, E. Fombonne, D. Geschwind, D. E. Grice, A. Klin, D. H. Ledbetter, C. Lord, C. L. Martin, D. M. Martin, R. Maxim, J. Miles, O. Ousley, B. Peterson, J. Piggot, C. Saulnier, M. W. State, W. Stone, J. S. Sutcliffe, C. A. Walsh and E. Wijsman) and the coordinators and staff at the SSC clinical sites; the SFARI staff, in particular N. Volfovsky; D. B. Goldstein for contributing to the experimental design; and the Rutgers University Cell and DNA repository for accessing biomaterials.

## Electronic MEdical Records and Genomics Network

Group Health Cooperative/University of Washington – Funding support for Alzheimer’s Disease Patient Registry (ADPR) and Adult Changes in Thought (ACT) study was provided by a U01 from the National Institute on Aging (Eric B. Larson, PI, U01AG006781). A gift from the 3M Corporation was used to expand the ACT cohort. DNA aliquots sufficient for GWAS from ADPR Probable AD cases, who had been enrolled in Genetic Differences in Alzheimer’s Cases and Controls (Walter Kukull, PI, R01 AG007584) and obtained under that grant, were made available to eMERGE without charge. Funding support for genotyping, which was performed at Johns Hopkins University, was provided by the NIH (U01HG004438). Genome-wide association analyses were supported through a Cooperative Agreement from the National Human Genome Research Institute, U01HG004610 (Eric B. Larson, PI).

Mayo Clinic – Samples and associated genotype and phenotype data used in this study were provided by the Mayo Clinic. Funding support for the Mayo Clinic was provided through a cooperative agreement with the National Human Genome Research Institute (NHGRI), Grant #: UOIHG004599; and by grant HL75794 from the National Heart Lung and Blood Institute (NHLBI). Funding support for genotyping, which was performed at The Broad Institute, was provided by the NIH (U01HG004424).

Marshfield Clinic Research Foundation – Funding support for the Personalized Medicine Research Project (PMRP) was provided through a cooperative agreement (U01HG004608) with the National Human Genome Research Institute (NHGRI), with additional funding from the National Institute for General Medical Sciences (NIGMS) The samples used for PMRP analyses were obtained with funding from Marshfield Clinic, Health Resources Service Administration Office of Rural Health Policy grant number D1A RH00025, and Wisconsin Department of Commerce Technology Development Fund contract number TDF FYO10718. Funding support for genotyping, which was performed at Johns Hopkins University, was provided by the NIH (U01HG004438).

Northwestern University – Samples and data used in this study were provided by the NUgene Project (www.nugene.org). Funding support for the NUgene Project was provided by the Northwestern University’s Center for Genetic Medicine, Northwestern University, and Northwestern Memorial Hospital. Assistance with phenotype harmonization was provided by the eMERGE Coordinating Center (Grant number U01HG04603). This study was funded through the NIH, NHGRI eMERGE Network (U01HG004609). Funding support for genotyping, which was performed at The Broad Institute, was provided by the NIH (U01HG004424).

Vanderbilt University - Funding support for the Vanderbilt Genome-Electronic Records (VGER) project was provided through a cooperative agreement (U01HG004603) with the National Human Genome Research Institute (NHGRI) with additional funding from the National Institute of General Medical Sciences (NIGMS). The dataset and samples used for the VGER analyses were obtained from Vanderbilt University Medical Center’s BioVU, which is supported by institutional funding and by the Vanderbilt CTSA grant UL1RR024975 from NCRR/NIH. Funding support for genotyping, which was performed at The Broad Institute, was provided by the NIH (U01HG004424).

Assistance with phenotype harmonization and genotype data cleaning was provided by the eMERGE Administrative Coordinating Center (U01HG004603) and the National Center for Biotechnology Information (NCBI). The datasets used for the analyses described in this manuscript were obtained from dbGaP at http://www.ncbi.nlm.nih.gov/gap through dbGaP accession number phs000360.v3.p1.

