## AdditionalFile.1 for "How rare and common risk variation jointly affect liability for autism spectrum disorder"

**Additional File 1**

| **Additional file 1: Table 1**. Treatment of potentially pathogenic variants from the PAGES subjects diagnosed with AD. “ASC 102” refers a variant falling into one of the 102 inferred ASD genes reported in Satterstrom et al.^10^ | | | |
| --- | --- | --- | --- |
|  | CNV | PTV (ASC 102) | MIS (ASC 102) |
| Initial count | 108 | 23 | 32 |
| Unique subjects | 96^1^ | 23 | 28 |
| After Selection^2^ | 88 | 20 | 23 |

^1^Mahjani et al. (in review) report one more carrier (16p11.2 deletion). This observation was removed during our QC.

^2^The data we analyze here are a subset of the data analyzed in Mahjani and were selected on the basis of European ancestry, same genotyping platform per case status, and quality control metrics on samples and genotypes. Our analyzed samples differ slightly from those in Mahjani because we emphasize different features of QC (i.e., European ancestry versus CNV calls for Mahjani.)

**Additional file 1: Table 2A.** Analysis of variance of GP as a function of ancestry cluster (CLS), diagnosis given cluster (DX|CLS), and eigenvectors (EVA).

| EVA | p-value for CLS | p-value for DX\|CLS |
| --- | --- | --- |
| 0^1^ | 0.497 | 7.61×10^-81^ |
| 0^2^ | 0.496 | 3.79×10^-79^ |
| 3^2^ | 0.536 | 9.93×10^-80^ |
| 10^2^ | 0.019 | 1.88×10^-68^ |

^1^ Based on the pair-out algorithm

^2^ Based on the one-out algorithm

**Additional file 1: Table 2B.** Logistic regression of ASD status on GP and carrier status on GP (cases only).

|  | ASD status | | | | Carrier status | | |
| --- | --- | --- | --- | --- | --- | --- | --- |
| EVA | OR | 95%CI | P | Pseudo-R^2^ (%) | OR | 95%CI | P |
| 0^1^ | 1.67 | 1.58-1.77 | 2.87×10^-74^ | 7.80 | 0.81 | 0.71-0.92 | 8.36×10^-4^ |
| 0^2^ | 1.66 | 1.57-1.76 | 7.67×10^-73^ | 7.64 | 0.81 | 0.71-0.92 | 8.56×10^-4^ |
| 3^2^ | 1.67 | 1.58-1.76 | 2.47×10^-73^ | 7.69 | 0.81 | 0.71-0.92 | 8.15×10^-4^ |
| 10^2^ | 1.60 | 1.51-1.69 | 1.16×10^-63^ | 6.60 | 0.81 | 0.72-0.91 | 3.87×10^-4^ |

^1^ Based on the pair-out algorithm

^2^ Based on the one-out algorithm

**Other approaches to normalizing allele frequencies**: Here we explore the effect of estimating allele frequency in other ways than that used in the main manuscript, the CLS; the other approaches are more typical of the methods described in the literature (Table S3). Two approaches were explored: estimating allele frequencies from the combined set of 3,011 affected and 3,011 genetically matched unaffected subjects (POP); and estimating the allele frequencies from the entire, combined sample of subjects (GCTA). Both approaches lead to a small bias in estimates of GP (Table S3). We also explore a variant of the GP estimation plan, rather than leaving out the matched pair and jointly estimating GP, we leave out only one subject and estimate GP, finding that this slight difference in sampling generates similar estimates of GP (Table s3).

**Additional file 1: Table 3.** Anova results of G-BLUP = CLS + DX when using different GRMs^1^

| GRM | P(CLS) | P(DX\|CLS) |
| --- | --- | --- |
| CLS^2^ | 0.496 | 7.61×10^-81^ |
| CLS^3^ | 0.496 | 3.79×10^-79^ |
| POP^3^ | 0.006 | 8.60×10^-79^ |
| GCTA^3^ | 0.006 | 1.72×10^-78^ |

^1^ Genomic prediction performed without adjusting for ancestry

^2^ Based on the pair-out algorithm

^3^ Based on the one-out algorithm

We next asked whether the effect of choosing different approaches to estimating population-level allele frequencies can be removed by using ancestry covariates (PCA) in the estimation model. For POP, we chose to use 3 EVA. For GCTA, we chose the standard 10 PCA, the number often used in analysis of genomic data when using the GCTA software (Table S4).

Using POP-GRM with 3 EVA results in a very strong CLS effect on GP and is therefore not recommended. Adding 10 PCA to the estimation model when using GCTA-GRM does control the effect of CLS on G-BLUP (p = 0.713). It does, however, come at a cost of losing predictive power of GP for ASD diagnosis status (OR = 1.54; 95%CI = 1.46-1.62; P = 1.59×10^-55^; pseudo-R^2^ = 5.68%) and carrier status (OR = 0.84; 95%CI = 0.74-0.95; P = 5.64×10^-3^).

**Additional file 1: Table 4.** Results from the Anova of G-BLUP = CLS + DX when using different GRMs but accounting for difference in ancestry by using different numbers of ancestry vectors.

| GRM | Ancestry Covariates | P(CLS) | P(DX\|CLS) |
| --- | --- | --- | --- |
| CLS^1^ | None | 0.496 | 7.61×10^-81^ |
| CLS^2^ | None | 0.496 | 3.79×10^-79^ |
| POP | 3 EVA | 1.63×10^-20^ | 1.01×10^-79^ |
| GCTA | 10 PCA | 0.713 | 5.55×10^-59^ |

**Training-Testing**

We used a pair-out (or single-out) approach to the training-testing algorithm, a computer intensive approach for calculation of the G-BLUP solutions for GP. A reasonable question is whether a more traditional and computationally less intensive training-testing – splitting data in larger subsets – would yield similar results. For this experiment, we randomly split the data in folds or splits and used a portion for training and the remainder for testing (Table S5-S6). Because these splits are random, we repeated this process 25 times. Notably, as the number of splits increases, accuracy approaches the two- or one-out approach, so careful selection of fold size could expedite calculations with only minor loss in accuracy.

**Additional file 1: Table 5.** Average results from the ANOVA of G-BLUP = CLS + DX when using different number of splits for of the data for training and testing (25 repetitions).

| Splits | P(CLS) | P(DX\|CLS) |
| --- | --- | --- |
| 2 | 0.298 | 1.10×10^-39^ |
| 4 | 0.365 | 8.79×10^-58^ |
| 10 | 0.487 | 1.42×10^-69^ |
| 20 | 0.449 | 1.15×10^-75^ |
| One-out | 0.496 | 3.79×10^-79^ |
| Pair-out | 0.497 | 7.61×10^-81^ |

**Additional file 1: Table 6.** Average results for the logistic regression of ASD status on G-BLUP and DMG status in cases on G-BLUP when using different splits of the data for training and testing 25 repetitions).

|  | ASD status | | | | DMG stats (cases only) | | |
| --- | --- | --- | --- | --- | --- | --- | --- |
| Splits | OR | 95%CI | P | Pseudo-R^2^ (%) | OR | 95%CI | P |
| 2 | 1.50 | 1.42-1.58 | 3.07×10^-38^ | 5.02 | 0.86 | 0.76-0.97 | 0.0379 |
| 4 | 1.58 | 1.50-1.67 | 1.51×10^-54^ | 6.35 | 0.83 | 0.74-0.94 | 0.0076 |
| 10 | 1.64 | 1.55-1.73 | 7.37×10^-65^ | 7.23 | 0.82 | 0.72-0.93 | 0.0021 |
| 20 | 1.66 | 1.57-1.75 | 5.21×10^-70^ | 7.60 | 0.82 | 0.72-0.92 | 0.0016 |
| One-out | 1.66 | 1.57-1.76 | 7.67×10^-73^ | 7.64 | 0.81 | 0.71-0.92 | 8.56×10^-4^ |
| Pair-out | 1.67 | 1.58-1.77 | 2.87×10^-74^ | 7.80 | 0.81 | 0.71-0.92 | 8.36×10^-4^ |

**Quality control for SNPs selected for PRS calculations**

SNPs from ASD and SCZ GWAS were selected using the following steps:

1. Select GWAS SNPs that were part of our imputed and QC-ed set of 5,145,175 SNPs.
2. Remove any SNP whose allele frequency in our samples deviates by more than 0.075 from the GWAS-estimated allele frequency.
3. Remove palindromic SNPs with minor allele frequency MAF>0.40.
4. Remove SNPs with MAF< 0.005.
5. Take only the most significant SNP in the MHC region. Remove all other SNP in the region chr6:25,000,000-34,000,000.
6. Clump the SNP using --clump in PLINK based on the p-values from the GWAS and the LD structure calculated from the control samples used for frequency calculations in our data (setting: clump-r^2^ = 0.50 and clump-kb = 50).
7. Use SNP with clump p-value < 0.01.

With SNPs selected by this procedure, PRS was calculated using --score in PLINK with option center. Centering of genotype counts was based on the frequencies in the controls that were not part of the matches in our data.

**Computational Efficiency**

For the general set-up of G-BLUP, see^20^. Solutions (GPs) can then obtained solving the equations Ax=b where x are the GPs. To expedite obtaining GPs, we first re-arrange A and b such that the matched pair of interest are in the last two rows and columns of A and the last two elements of b. We can then solve for this pair as follows. Let the Cholesky decomposition of A be L and A=LL’, in which L is a lower triangular matrix. Now, to solve Ax=b, first solve Lz=b where z=L’x. This is the forward step. Because of the special structure of L, these solutions can be obtained using row-wise elimination avoiding calculation of the inverse of L. Once z has been obtained, solve L’x=z to obtain the solutions x using the elimination in reverse, the backward step. Because we are only interested in the solutions for the last two equations, the pairs whose GP are to be predicted, the backward step only involves solving the last two equations, which we do through backwards elimination.

**Additional file 1: Figure 1.** Projection of genetic ancestry on the first two dimensions of the spectral decomposition. The PAGES data we analyze here are a subset of the data analyzed in Mahjani et al. (in review) and were selected on the basis of European ancestry, same genotyping platform per case status, and quality control metrics.

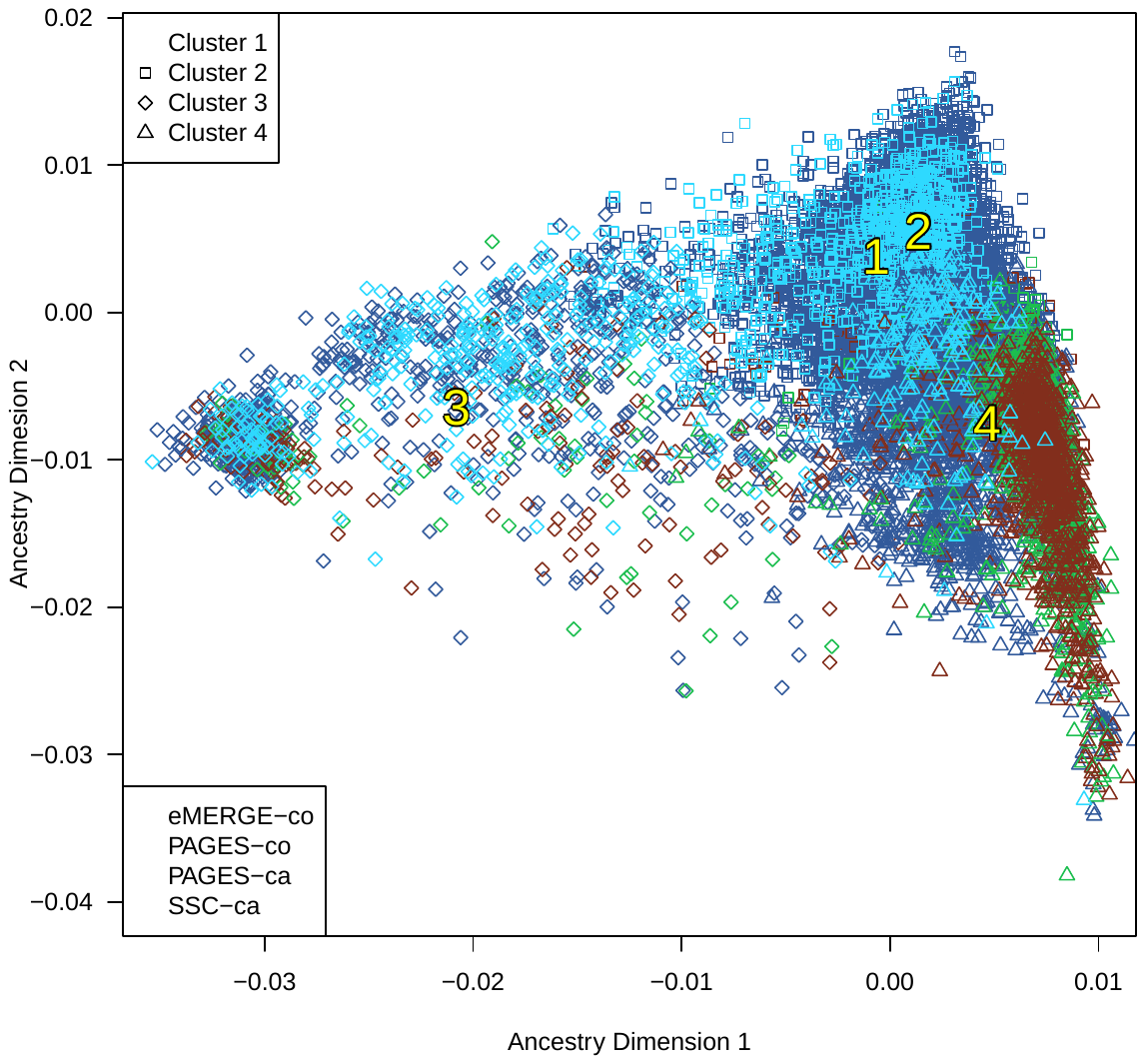

**Additional file 1: Figure 2**. Case-control balance. Results of 100 simulations of different degrees of imbalance and showing the distribution of p-values for a cluster effect on burden. (A) A balanced, *but not matched* design of 300 cases and 300 controls for each of four ancestry clusters; (B) An unbalanced design in which 300 ASD subjects per cluster were contrasted with a varying, but unbalanced number of unaffected subjects (330, 315, 285, or 270 unaffected subjects, randomly assigned per cluster and per simulation); (C) The unbalanced design of (B), but using three eigenvectors of ancestry to account for differences among ASD and unaffected subjects.

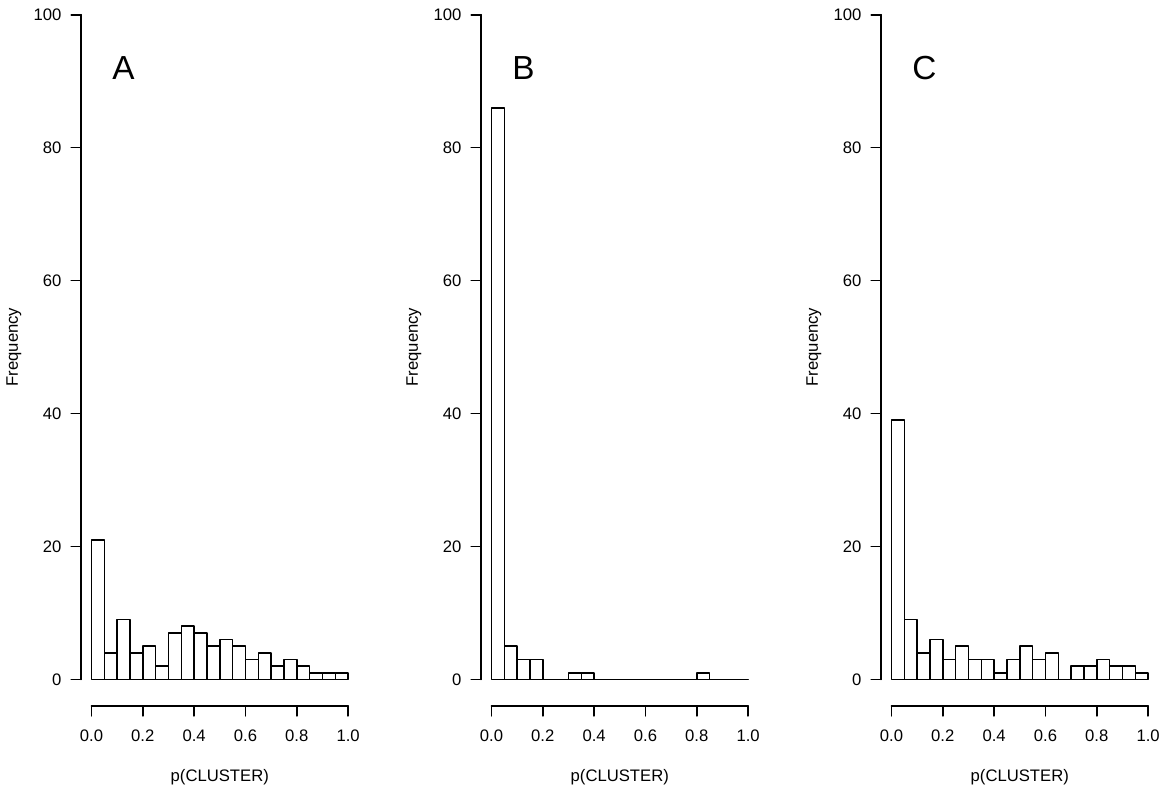
